# A quantitative measure of choroid plexus contrast enhancement strongly relates to markers of diffuse brain tissue injury in multiple sclerosis

**DOI:** 10.1101/2025.03.26.25324694

**Authors:** Sneha Senthil, Ian Tagge, Dumitru Fetco, Cheng Hsun Hsieh, Haz-Edine Assemlal, Zahra Karimaghaloo, Emily Fetco, G.R. Wayne Moore, Douglas L. Arnold, David A. Rudko, Sridar Narayanan

## Abstract

**Objective:** Recent studies suggest that disruptions of the blood–cerebrospinal fluid barrier within the choroid plexus (ChP) may contribute to MS pathogenesis. We investigated the relationship between a quantitative marker of ChP enhancement and markers of focal and diffuse brain tissue injury in multiple sclerosis (MS).

**Methods:** 34 MS participants underwent 7T MRI including MP2RAGE-based qT1 mapping pre- and post-contrast, and FLAIR acquisitions. “Delta T1” (ΔT1) maps were calculated by subtraction of post-contrast from co-registered pre-contrast qT1 maps. ChP, white matter lesions (WML), normal-appearing white matter (NAWM) and grey matter (GM) were segmented. Linear regression analyses were conducted between mean ΔT1 values of ChP and (1) WML volume, (2) pre-Gd mean qT1 of WML, (3) pre-Gd mean qT1 of NAWM, and (4) pre-Gd mean qT1 of GM.

**Results:** ΔT1 of ChP was significantly associated with pre-Gd qT1 of NAWM (β =0.20, R^2^ = 0.54, p<0.001) and GM (β = 0.32, R^2^ = 0.62, p<0.001). No significant associations were found between ChP ΔT1 and WML volume (p = 0.3) or WML qT1 (p = 0.05).

**Interpretation:** The strong associations we observed between the degree of ChP contrast enhancement and markers of diffuse brain tissue injury, combined with a lack of a relationship with lesion volume or qT1 within lesions, support the hypothesis that entry of toxic factors into the CSF via the ChP may constitute an additional mechanism of brain tissue injury distinct from the classic lesion-based pathology of MS.

## Introduction

Multiple sclerosis (MS) is a chronic demyelinating disease that results in a wide range of neurological deficits. While immune cell trafficking into the central nervous system (CNS) through disruptions in the blood-brain barrier (BBB) is a well-established feature of MS (1–4), recent studies have implicated the choroid plexus (ChP) in MS pathogenesis (5–7). The ChP, a highly vascularized structure within the brain’s ventricular system, serves as an entry point for lymphocytes and plays a pivotal role in modulating inflammatory processes (5,8,9).

In addition to its immune functions, the ChP plays a vital role in the production of cerebrospinal fluid (CSF), which is essential for maintaining the homeostasis of the CNS. It forms the blood-CSF barrier (BCSFB), a selective interface of epithelial tight junctions (6), and harbors immune cells that protect against pathogens and regulate immune surveillance via chemokine signaling (8). Disruption of the BCSFB, however, can compromise CNS defense by increasing ChP permeability, allowing harmful microbes to infiltrate into the CSF (10). In inflammatory conditions, alterations in ChP permeability allows pathogenic T-lymphocytes to migrate through both the BBB and the BCSFB, contributing to increased infiltration of immune cells and pro-inflammatory factors into the CNS (11). Given the central role of neuroinflammation in MS, where pathological changes like axonal loss, demyelination, and neurodegeneration collectively drive disease severity, investigating the contribution of increased BCSFB permeability to brain tissue injury is crucial, yet remains largely unexplored.

Quantitative T1 (qT1; longitudinal relaxation time constant) mapping on MRI offers non-invasive assessment of brain tissue microstructure. MRI-histology studies have shown that prolonged qT1 reflects increased water content, demyelination, and axonal loss in white matter (WM), serving as a marker of tissue injury (12–14). Previous studies have also demonstrated that qT1 correlates with tissue damage in MS, with elevated qT1 observed in WM lesions (WMLs) compared to normal-appearing white matter (NAWM) (15) and in NAWM (16–22) and normal-appearing gray matter (NAGM) (18,23,24) compared to healthy controls. Recently, the Magnetization Prepared 2 Rapid Acquisition Gradient Echoes (MP2RAGE) technique has been employed for qT1 measurement, providing both T1-weighted MPRAGE-like images and qT1 maps (25). MP2RAGE can be acquired at high spatial resolution within clinically acceptable scan times, and is self-bias-field corrected, making it well-suited for ultra-high-field applications (>=7T) (25). It has been demonstrated that a change in qT1 in tissue after administration of a gadolinium (Gd)-based contrast agent (GBCA) can be measured through voxel-wise subtraction of qT1 maps pre- and post-Gd (26). ΔT1, the difference in qT1 pre- and post-Gd, could also provide insight into the permeability of the BCSFB.

We aimed to use qT1 mapping to assess the relationship of T1 shortening (ΔT1) in the ChP after intravenous Gd-contrast administration, with markers of focal and diffuse brain tissue injury, namely, WML volume, qT1 within WML, and qT1 in NAWM, and grey matter (GM). We hypothesize that greater ChP enhancement (higher ΔT1) correlates with greater tissue damage in MS participants, supporting the idea that systemically circulating molecules and/or inflammatory cells entering the CNS through the ChP and diffusing throughout the brain via the CSF is a mechanism of tissue injury in MS.

## Methods

### Study Participants

We recruited 34 people with MS (RRMS = 24, SPMS = 10; age = 53.2 ± 9.8; M= 7, F= 27, EDSS [Expanded Disability Status Scale] = 3.1 ± 1.8, disease duration = 22.6 ± 8.6 years), followed at the MS Clinic of the Montreal Neurological Institute-Hospital. Inclusion criteria included an MS diagnosis based on the 2017 revised McDonald criteria (27), age ≥ 18 years, and being either untreated or on a stable disease modifying therapy (DMT) for at least six months (i.e. without a recent switch in DMT). Exclusion criteria were: concomitant neurological disease in addition to MS, relapse or treatment with steroids within 30 days of the baseline visit, a history of claustrophobia, and contraindications to MRI, such as cardiac pacemakers, prosthetic heart valves, cochlear implants, or any metallic implants in the body. Detailed demographic information is presented in Table 1. All study procedures were approved by the hospital’s Research Ethics Board and conducted in accordance with the Declaration of Helsinki. Written informed consent was obtained from all study participants.

**Table 1:**
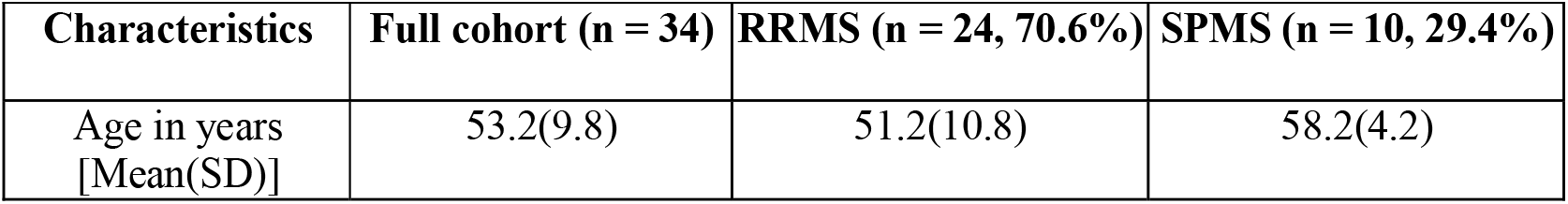
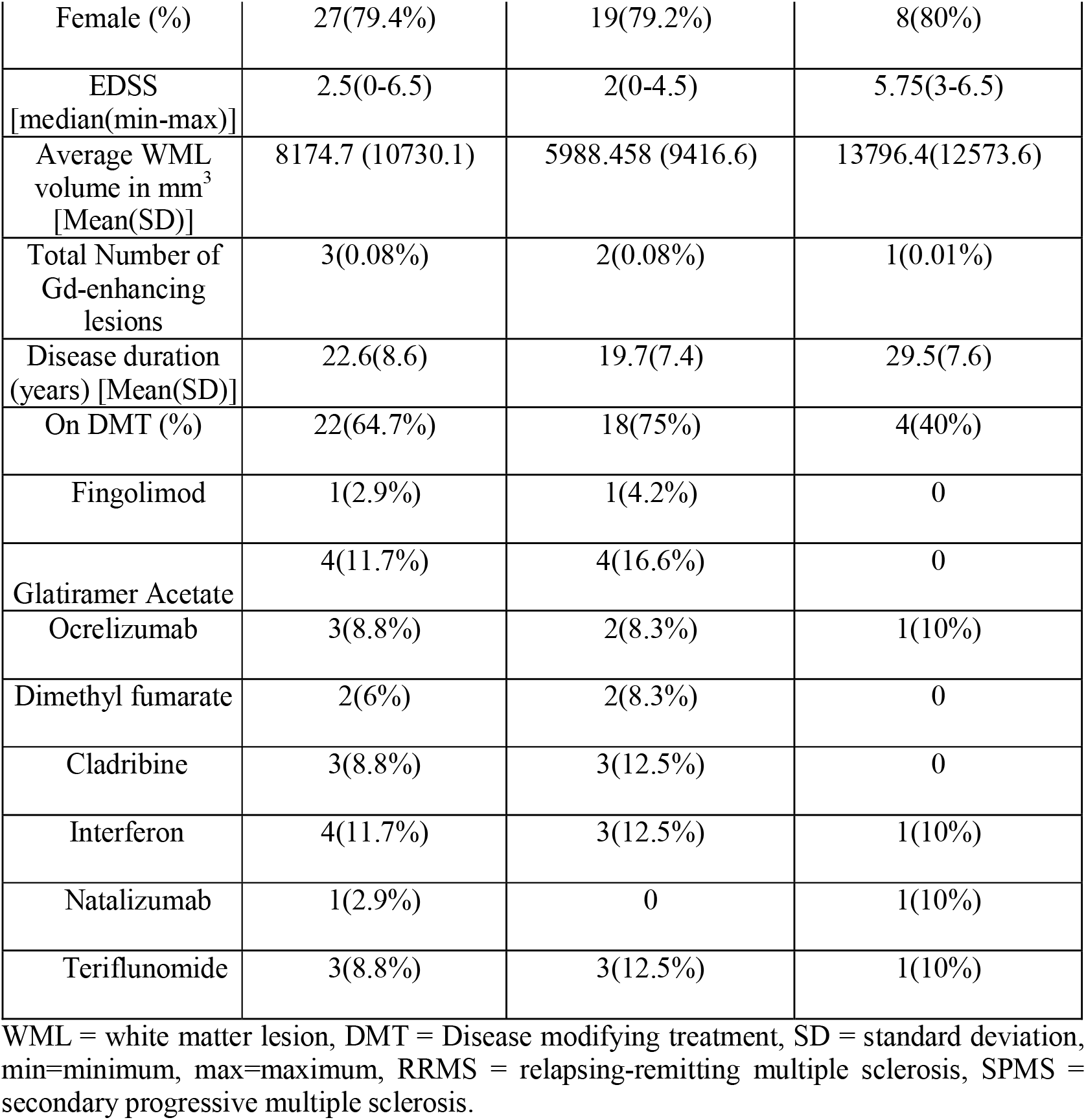
Detailed demographic information of the recruited participant cohort.

| Characteristics | Full cohort (n = 34) | RRMS (n = 24, 70.6%) | SPMS (n = 10, 29.4%) |
| --- | --- | --- | --- |
| Age in years<br>[Mean(SD)] | 53.2(9.8) | 51.2(10.8) | 58.2(4.2) |
| Female (%) | 27(79.4%) | 19(79.2%) | 8(80%) |
| EDSS<br>[median(min-max)] | 2.5(0-6.5) | 2(0-4.5) | 5.75(3-6.5) |
| Average WML<br>volume in mm<sup>3</sup><br>[Mean(SD)] | 8174.7 (10730.1) | 5988.458 (9416.6) | 13796.4(12573.6) |
| Total Number of<br>Gd-enhancing<br>lesions | 3(0.08%) | 2(0.08%) | 1(0.01%) |
| Disease duration<br>(years) [Mean(SD)] | 22.6(8.6) | 19.7(7.4) | 29.5(7.6) |
| On DMT (%) | 22(64.7%) | 18(75%) | 4(40%) |
| Fingolimod | 1(2.9%) | 1(4.2%) | 0 |
| Glatiramer Acetate | 4(11.7%) | 4(16.6%) | 0 |
| Ocrelizumab | 3(8.8%) | 2(8.3%) | 1(10%) |
| Dimethyl fumarate | 2(6%) | 2(8.3%) | 0 |
| Cladribine | 3(8.8%) | 3(12.5%) | 0 |
| Interferon | 4(11.7%) | 3(12.5%) | 1(10%) |
| Natalizumab | 1(2.9%) | 0 | 1(10%) |
| Teriflunomide | 3(8.8%) | 3(12.5%) | 1(10%) |

WML = white matter lesion, DMT = Disease modifying treatment, SD = standard deviation, min=minimum, max=maximum, RRMS = relapsing-remitting multiple sclerosis, SPMS = secondary progressive multiple sclerosis.

### MRI acquisition

Participants were scanned on a 7T whole-body MR scanner (MAGNETOM Terra, Siemens Healthineers, Erlangen, Germany) with an 8 Tx/32 Rx-channel head coil (Nova Medical Inc., Wakefield, MA, USA) in parallel transmission (pTx) mode. The imaging protocol included an MP2RAGE acquisition, both pre- and 8 minutes post-Gd injection, with the following sequence parameters: resolution = 0.7mm^3^ isotropic, FOV = 240×240×172 mm^3^, TI1/TI2 = 800 ms/2700 ms, TR/TE = 6000 ms/2.74 ms, acquisition time = 10:14 min. A T1 map was computed using the Siemens MapIt software for each MP2RAGE acquisition. A 3D T2-weighted Fluid Attenuated Inversion Recovery (FLAIR) sequence was acquired 20 minutes post-Gd injection with the following sequence parameters: resolution = 0.7mm^3^ isotropic, FOV = 225×225×179.2 mm^3^, TR/TE = 10560 ms/301 ms, TI1 = 2500 ms, acquisition time= 9:32. Visual inspection of all images on the scanner console confirmed that no severe motion artifacts were present that could impact image processing.

### Image processing

All images were processed using the MINC toolkit (https://bic-mni.github.io/)(28). All MRI images for each participant were individually co-registered to their native FLAIR space to ensure spatial alignment. A brain mask was generated from the target modality using a U-net-based deep learning segmentation method (29), which was then resampled and applied to all MRI images. Intensity inhomogeneities in the FLAIR images were corrected by the N4 bias field algorithm (30). “Delta T1” (ΔT1) maps were calculated by voxel-by-voxel subtraction of post-contrast from pre-contrast T1 maps of each participant.

### Tissue and Lesion segmentations

Manual segmentation of T2-hyperintense WMLs (on FLAIR images) (author D.F) and ChP (on pre-Gd-T1-weighted images) (authors S.S, C.H.H, E.F) was performed using the Display software (http://www.bic.mni.mcgill.ca/software/Display). An experienced medical image analyst (author D.F, >20 years of experience) carefully reviewed and corrected all segmentation masks, and WML volume was calculated for each participant using the MINC tools.

Tissue segmentation was performed using the Brain Tissue Composition-Net (BTC-Net) pipeline developed at NeuroRx. This pipeline involves generating tissue segmentation masks using a custom 3D convolutional neural network, inspired by the U-Net architecture (29). The model was trained on an internal dataset of co-registered MRI scans with a 1 mm^3^ isotropic resolution, comprising 130 participant sessions. The ground truth for healthy tissue segmentation was established using an automated multi-atlas label fusion method (31), with lesion masks manually reviewed and corrected. The generated WM and GM (inclusive of deep grey and cortical grey matter) masks excluded the cerebellum due to its complex structure and decreased segmentation reliability owing to partial volume effects. NAWM masks were generated by explicitly masking out the WM lesion mask from the BTC-Net generated tissue masks. The resulting NAWM and GM tissue masks were eroded by 0.7mm in each direction (i.e., 1 voxel) to minimize the inclusion of border voxels with partial volume. A representative example of tissue and lesion segmentations is illustrated in Figure 1. Mean ΔT1 of ChP was calculated by applying the ChP segmentation mask to the ΔT1 maps for each participant. Mean pre-Gd qT1 of GM, NAWM, and WM lesions were obtained by applying the respective masks to the pre-Gd qT1 map.

**Figure 1:**
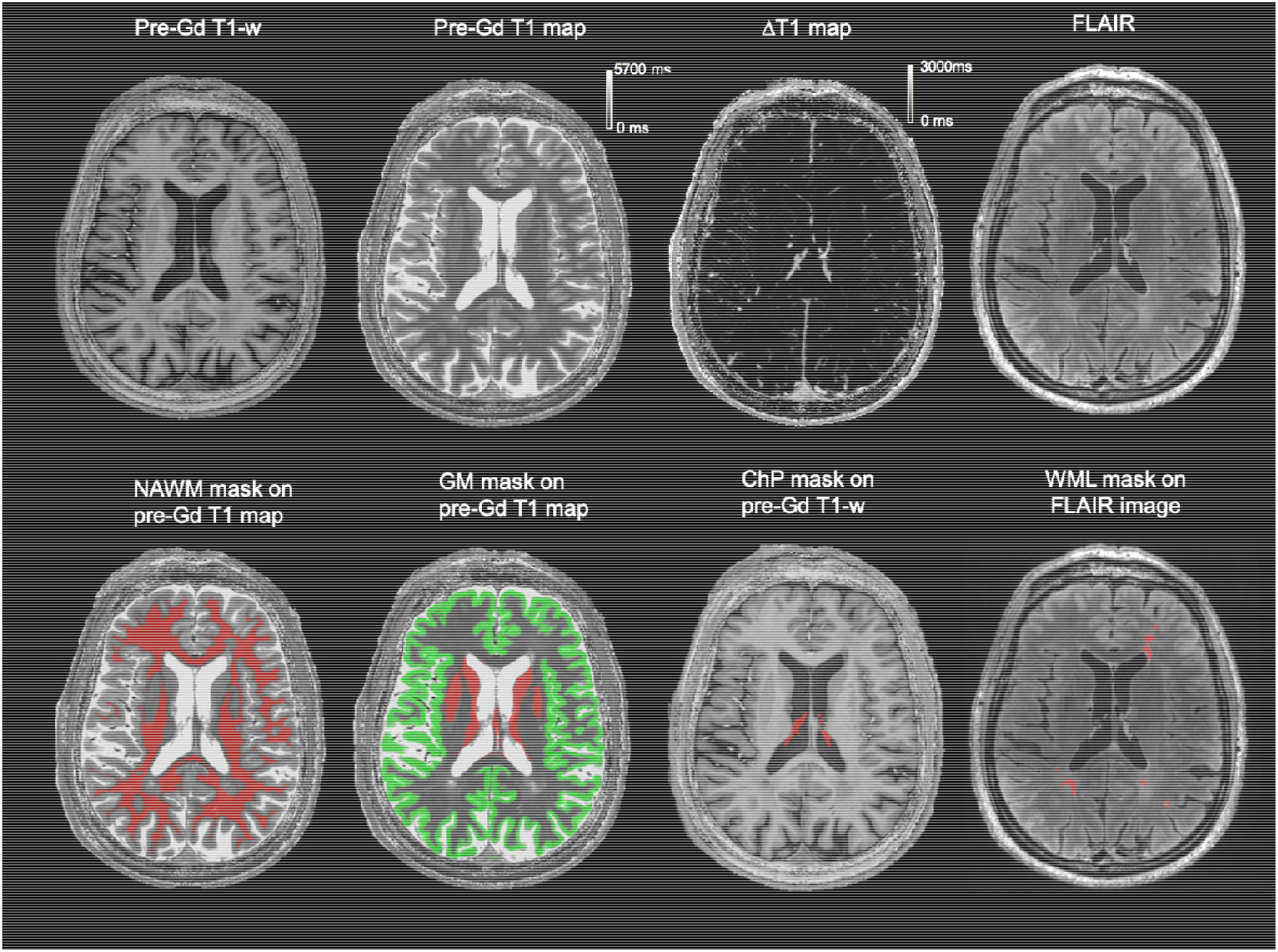
Representative tissue and lesion segmentations for an RRMS participant included in our study. The segmentations are co-registered to the participant’s native 7T FLAIR image space. Top row displays pre-Gd T1-w image; pre-Gd T1 map; ΔT1 map; FLAIR image (left to right). The bottom row displays the segmentation masks for NAWM, GM (deep GM in red and cortical GM in green), ChP, and WML (left to right).

### Statistical analysis

All statistical analyses were performed using R (v4.4.0; R Core Team, 2024). Differences between qT1 measures of WML and NAWM were assessed using an unpaired two-tailed Student’s t-test. In alignment with our hypothesis, separate linear regression analyses were carried out between mean ΔT1 values of ChP as the predictor variable and the following response variables: 1) WML volume; 2) pre-Gd mean qT1 values of WML; 3) pre-Gd mean qT1 values of NAWM; and 4) pre-Gd mean qT1 values of GM. The ChP volume of each subject was included as a covariate in these linear models, as smaller ChP ROIs may have resulted in increased partial volume effects, potentially biasing the measured T1 change. Additionally, Pearson correlation coefficients were determined to assess the strength of each of these relationships, with 95% confidence intervals provided for each analysis. To address potential outliers in the qT1 data, a 1.5 interquartile range (IQR) filter was applied to the data, excluding values that fell outside 1.5 times the IQR from the first and third quartiles. Data points identified as outliers were removed from further analysis. To examine the relationship between ΔT1 of ChP and disease severity, as assessed by EDSS scores, a Spearman’s rank correlation was conducted. Statistical significance was evaluated using a significance level of α=0.05. *P* values were Bonferroni-corrected for multiple comparisons (α = 0.05/5=0.01).

## Results

### Study population

Demographic and clinical characteristics of the recruited cohort are shown in Table 1. The MS cohort included 24 participants with RRMS (70.6%) and 10 participants with SPMS (30.4%). Most participants (64.7%) were on disease-modifying therapies (DMTs) at the time of their scan. Visual assessment of post-contrast FLAIR and T1-weighted images by two neurologists revealed only 3 (9%) participants with Gd-enhancing lesions.

### Data Analysis

Table 2 summarizes the data obtained from ΔT1 and qT1 maps. Consistent with previous literature (23,32), pre-Gd qT1 in WML was significantly higher than that of NAWM (unpaired t-test, p<0.001). No significant differences were seen between pre-Gd qT1 of lesions of RRMS and SPMS participants.

**Table 2:** Summary of T1 and ΔT1 metrics obtained from segmentation masks used for linear regression analyses. NAWM and GM masks were eroded by one voxel. To get a positive measure of ChP enhancement, the ΔT1 map was obtained by subtracting the post-qT1 maps from the pre-qT1 maps.

| Segmented Mask | Measure | Mean (SD) | Median [min, max] |
| --- | --- | --- | --- |
| ChP | $\Delta T1$ (ms) | 1629.9 (225.2) | 1595.5 [1334.6, 2159.74] |
|  | qT1 pre-Gd (ms) | 2860.9 (326.5) | 2824.8 [2047.4, 3581.1] |
|  | qT1 post-Gd (ms) | 1281.2 (177.7) | 1286.12 [910.2, 1734.5] |
|  | Volume (mm<sup>3</sup>) | 1439.1 (597.7) | 1377.2 [626.9, 2906.2] |
| WML | $\Delta T1$ (ms) | 86.7 (28.7 ) | 73.2 [47.4,155] |
|  | qT1 pre-Gd (ms) | 1974.2 (275.9) | 1917.4 [1514.4, 2768.1] |
|  | qT1 post-Gd (ms) | 1861.8 (281.1) | 1832.9 [1247.65, 2607.8] |
|  | Volume (mm<sup>3</sup>) | 9222.1 (10538.7) | 5369.0 [553.3, 40700.7] |
| NAWM | $\Delta T1$ (ms) | 32.0 (6.6) | 31.1 [18.8, 48.7] |
|  | qT1 pre-Gd (ms) | 1244.5(115.2) | 1212.9 [1094.5, 1584.4] |
|  | qT1 post-Gd (ms) | 1225.7 (113.0) | 1212.6 [1074.8, 1551] |
| GM<br>(consisting of<br>deep and<br>cortical GM) | $\Delta T1$ (ms) | 176.7 (41.5) | 170.6 [86.6, 279.9.0] |
|  | qT1 pre-Gd (ms) | 1958.1(210.5) | 1844.3 [1720.3, 2303.1] |
|  | qT1 post-Gd (ms) | 1755.5(174.6) | 1675.9 [1445.6, 2090.4] |
| Deep GM | $\Delta T1$ (ms) | 74.36 (138.08) | 117.07 [-160.53,484.86] |
|  | qT1 pre-Gd (ms) | 1959.0(226.4) | 1953.6 [1554.3, 2371.8] |
|  | qT1 post-Gd (ms) | 1755.5(174.6) | 1675.9 [1445.6, 2090.4] |
| Cortical GM | $\Delta T1$ (ms) | 195.38 (105.20) | 207.97 [-246.98, 346.08] |
|  | qT1 pre-Gd (ms) | 1987.7(217.5) | 1866.5 [1745.2, 2354.5] |
|  | qT1 post-Gd<br>(ms) | 1755.5(174.6) | 1675.9 [1445.6, 2090.4] |

The results of separate linear regression analyses evaluating the relationships between mean ΔT1 values of ChP and markers of brain tissue injury are shown in Figure 2. We observed significant predictive relationships between ΔT1 values in ChP and pre-Gd qT1 values of both NAWM (β =0.20, R^2^ = 0.54, F(2, 28) = 16.21, p<0.001) and GM (β = 0.32, R^2^ = 0.62, F(2, 28) = 23.03, p<0.001). No significant association of ΔT1 values of ChP with WML volume (β =-6.12, *p*=0.3) or pre-Gd qT1 of WML (β =0.31, *p*=0.05) was seen. Pearson correlation analyses showed a strong positive association between ΔT1 of ChP and pre-Gd qT1 of NAWM (*r* = 0.73, *p* < 0.001) and pre-Gd qT1 of GM (*r* = 0.71, *p* < 0.001). In contrast, neither the correlation between ΔT1 of ChP and pre-Gd qT1 of WML (*r*=0.48, *p*>0.05) nor the correlation between ΔT1 of ChP and WML volume (r=-0.24, p>0.05) were significant. Furthermore, no significant correlation was observed between ΔT1 of ChP and EDSS (Spearman’s ρ = 0.04, p = 0.822).

**Figure 2:**
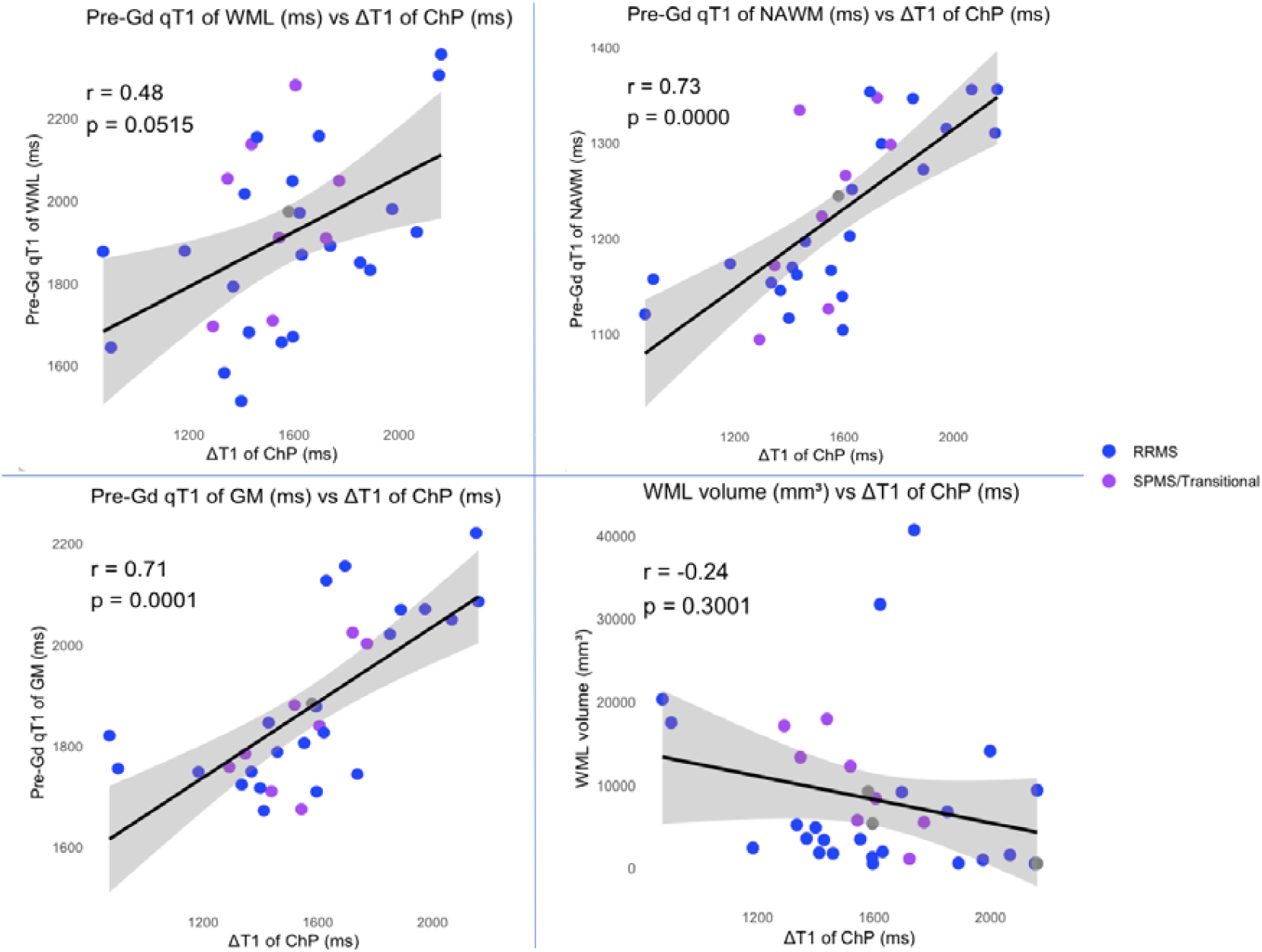
Shows the scatterplots between ΔT1 of ChP and (A) pre-Gd mean qT1 values of WML (*r=0*.*48*,*p>0*.*05*) (B) pre-Gd mean qT1 values of NAWM (*r=0*.*73*,*p<0*.*001*), (C) pre-Gd mean qT1 values of GM (*r=0*.*71*,*p<0*.*001*) and (D) WML volume (*r=-0*.*24*,*p>0*.*05*). The r and p-values of the whole linear regression model are reported on the top left corner of each graph. The shaded region shows 95% confidence intervals.

## Discussion

In this study, we applied qT1 mapping to assess the relationship between the degree of ChP enhancement after GBCA injection, our proposed marker of BCSF barrier permeability, and markers of brain tissue injury in MS. Pre-Gd qT1 values were used to assess brain tissue injury as they reflect tissue characteristics unaffected by the dynamics of Gd contrast enhancement. We found that contrast-related T1-shortening (ΔT1) of the ChP was a significant predictor of mean pre-Gd qT1 of NAWM and GM. Pearson correlation analyses revealed strong positive associations between ΔT1 of ChP and pre-Gd qT1 of NAWM and GM, thereby supporting the hypothesis that the degree of impairment of the BCSFB in MS is related quantitatively to the degree of diffuse brain tissue injury in normal-appearing tissues. In contrast, we did not observe significant correlations of ΔT1 of the ChP with WML volume nor with pre-Gd qT1 values within WML, where the overt pathology associated with BBB breakdown and focal lesion formation is expected to predominate over more diffuse pathology (33). Previous studies using histopathological analyses have revealed that WMLs primarily arise due to localized immune cell infiltration through a compromised BBB, enabling perivascular inflammation, microglial activation, and demyelination (34). Unlike the BCSFB, which regulates CSF composition and contributes to global tissue injury, the BBB is a highly selective barrier that, when disrupted, allows the entry of circulating immune cells, such as T cells and B cells, into the CNS parenchyma (35). This infiltration triggers a cascade of inflammatory processes, including oxidative stress, fibrinogen leakage, complement activation, and sustained microglial activation, all of which drive lesion formation. However, it is also important to consider that WMLs represent a more advanced stage of pathology than NAWM and their initiation may have originally involved similar changes to those seen in NAWM, potentially influenced by ChP permeability. Prior to lesion formation, the tissue in these regions may have undergone injury similar to NAWM, suggesting that BCSFB dysfunction could have played a role in the early stages of lesion development. This would be consistent with our prior findings that regions of NAWM destined to become lesions exhibit abnormal estimates of myelin volume fraction and axon volume fraction on quantitative MRI (36). Given that there is bulk flow of CSF through the brain parenchyma, toxic factors released into the CSF may contribute not only to diffuse pathology of the NAWM, but also plant seeds of injury that later result in the formation of focal lesions (37). In future, it would be valuable to explore the relationship between markers of BCSFB permeability with *surface-in* gradients of damage (periventricular and subpial). Such surface-in gradients are more likely to be affected by CSF-driven inflammatory processes linked to BCSFB dysfunction (38).

In order to contextualize our findings, it is essential to explore the potential underlying mechanisms by which the BCSFB integrity can be compromised. The BCSFB is characterized by the presence of fenestrated capillaries within the ChP, where the true “barrier” function is performed by the ChP epithelium. This barrier is formed by tight junctions between adjacent epithelial cells, composed of key proteins such as occludin, claudins (including claudin-1, -2, -3, and -11), and zona occludens-1, which regulate selective permeability and maintain the integrity of the blood-CSF interface (39). Among these, claudin-2 in tight junctions plays a crucial role in modulating permeability at the BCSFB, which is inherently more permeable than the BBB (40). During periods of inflammation, claudin-2 expression is upregulated (41), increasing paracellular permeability and allowing the passage of cytokines, immune cells, and proteins into the CSF. This upregulation is driven by pro-inflammatory cytokines such as Interleukin 6 (IL-6), Tumor Necrosis Factor-alpha (TNF-α), and Interferon-gamma (IFN-γ), which stimulate claudin-2 expression, further weakening barrier function (42). Concurrently, the downregulation or mislocalization of other tight junction proteins, such as claudin-11, may further disrupt the BCSFB, weakening its ability to regulate CSF composition and maintain CNS homeostasis (43).

Given evidence of BCSFB breakdown, qT1-mapping was used to derive ΔT1 values in ChP on Gd-enhanced MRI, our proposed BCSFB permeability marker. The endothelial fenestrae in the ChP allow the passive diffusion of certain molecules (up to ~70 kDa), facilitating communication between the CNS and peripheral circulation under steady-state conditions (44). Molecules larger than this threshold generally remain confined to the vascular lumen, preventing their entry into the ChP stroma. The GBCA used in this study, gadobutrol (604.7 Da), is small enough to pass through ChP endothelial fenestrations and into the underlying stroma, contributing to measurable T1 shortening (44,45). As for pre-Gd qT1 measurements, the calculated mean values in NAWM and GM across all participants were 1356.17 ± 132.69ms and 2553.59 ± 365.90ms respectively. These values are in concordance with previously published ranges at 7T (46). Values of pre-Gd qT1 in WM lesions (1974.2 ± 275.9 ms) were significantly prolonged [p<0.001], consistent with previous studies (23,32), indicating microstructural damage within WMLs, likely driven by myelin degradation, with potential contributions from axonal loss and increased tissue water content due to neuroinflammation(7) or edema (47).

Several other quantitative imaging techniques like magnetization transfer (MT) imaging, diffusion tensor imaging (DTI), T2* mapping and dynamic contrast-enhanced (DCE) MRI could further refine our assessment of BCSFB integrity and its relation to brain tissue injury. Specifically, quantitative measurements of vascular permeability using DCE-MRI, which has been used widely to study BBB breakdown in NAWM (26), may provide important dynamic measures of contrast agent extravasation into the ventricular CSF (48).

There are several limitations in this study that should be considered when evaluating the conclusions of this work. From a technical standpoint, the inversion times of our MP2RAGE acquisition were optimized for maximising GM-WM contrast and the calculation of qT1 in brain parenchyma, and were not ideal for assessing qT1 or qT1 change in CSF, which has a very long T1. Thus we were not able to assess qT1 reliably in the ventricles themselves. The absence of contrast-enhanced MRI of healthy participants due to ethical concerns precluded comparison of the distribution of ΔT1 of ChP in MS participants to that in healthy people. However, the strong association of ΔT1 in ChP with markers of brain tissue injury suggest that the degree of ChP enhancement we observed in MS participants is beyond that expected in healthy people. Our sample size was relatively small and weighted toward people with RRMS, limiting generalizability of the results. However, the strength of the associations we observed between the degree of ChP enhancement and markers of brain tissue injury increases confidence that our findings are robust. The cross sectional nature of the study limits us from exploring how our putative marker of BCSFB permeability changes with time and relates to disease progression. Future longitudinal studies with larger sample sizes and greater representation of people with progressive MS are warranted.

## Conclusion

The strong associations we observed between the degree of ChP contrast enhancement and markers of diffuse brain tissue injury, combined with a lack of a relationship with lesion volume or qT1 within lesions, support the hypothesis that entry of toxic factors into the CSF via the ChP may constitute an additional mechanism of brain tissue injury distinct from the classic lesion-based pathology of MS.

## Data Availability

The data that support the findings of this study are not publicly available due to concerns surrounding patient confidentiality but are available from the corresponding author on reasonable request.

## Acknowledgements

This study was supported in part by the Canadian Institutes of Health Research, grant #153005 (SN), the United States Department of Defense, Multiple Sclerosis Research Program, Investigator-Initiated Research Award (Award No. W81XWH19-1-0486) (DAR) and an Investigator-Initiated award from Novartis Canada (DLA). SS thanks the Fonds de Recherche Québec – Santé for personal support (scholarship). The authors are grateful to all the participants who made this study possible. We also thank Rozie Arnaoutelis for invaluable help in project coordination, Risavarshini Thevakumaran for helpful discussions, and our MRI technologists Ronaldo Lopez, David Costa and Soheil Mollamohseni Quchani for their outstanding work.

## Author Contributions

S.N., D.A.R. and S.S. contributed to conception and design of the manuscript; S.S., S.N., D.A.R., I.T., D.F., C.H.H., H.E.A., Z.K. and E.F. contributed to acquisition, processing and analysis of data; S.S., G.R.W.M., D.A.R., D.L.A. and S.N. contributed to drafting the text and preparing figures. All authors approved the manuscript.

